# Monitoring SARS-CoV-2 Populations in Wastewater by Amplicon Sequencing and Using the Novel Program SAM Refiner

**DOI:** 10.1101/2021.06.24.21259469

**Authors:** Devon A. Gregory, Chris G. Wieberg, Jeff Wenzel, Chung-Ho Lin, Marc C. Johnson

## Abstract

Sequencing SARS-CoV-2 from wastewater has become a useful tool in monitoring the spread of variants. We use a novel computation workflow with SARS-CoV-2 amplicon sequencing in order to track wastewater populations of the virus. As part of this workflow, we developed a program for both variant reporting and removal of PCR generated chimeric sequences. With these methods, we are able to track viral population dynamics over time. We observe the emergence of the variants of concern B.1.1.7 and P.1, and their displacement of the D614G B.1 variant.

## 1. Introduction

SARS-CoV-2 became pandemic and caused a world-wide health crisis starting in 2020 [1]. Full genome sequences of SARS-CoV-2 were rapidly made available within the first months of spread [2, 3]. Partial and whole genome sequencing of SARS-CoV-2 has been an important tool in monitoring transmission paths and the emergence of variant lineages. Most sequencing of SARS-CoV-2 has been done on clinical samples. However, early in the SARS-CoV-2 pandemic, wastewater began to be used to track community levels and spread of SARS-CoV-2 by RT-qPCR methods [4, 5]. Investigators have also used high throughput sequencing on wastewater samples to obtain full or partial SARS-CoV-2 genomic sequences which were used for metagenomic and epidemiologic analysis [6, 7, 8, 9, 10, 11, 12, 13]. Sequences identified in wastewater samples may reflect known lineages, as well as lineages not reported from clinical samples. Combinations of mutations not observed in clinical samples may represent new infections not yet picked up by clinical sampling or lineages that are under-represented in clinical samples. Approaches using wastewater are particularly relevant with the emergence of variant lineages that may vary from previous isolates in their fitness and/or disease.

The state of Missouri has been monitoring wastewater with RT-qPCR to track the prevalence and spread of SARS-CoV-2 (https://storymaps.arcgis.com/stories/f7f5492486114da6b5d6fdc07f81aacf). We sought to begin using the same samples for high throughput sequencing to track the presence and spread of known and previously unreported variant lineages. We were specifically interested in the spike gene and used primers to target 3 regions for amplification, the N-terminal domain (NTD), receptor binding domain (RBD) and the region of the S1 and S2 subunit split (S1S2). We chose these regions due to the numerous variations matching evolving lineages found in them and their significance in potential immune evasion [14]. While there are a number of high throughput sequencing technologies and methods, the sequence output is relatively standard; the processing and analysis of that sequence data is not. There are numerous programs and pipelines that can be used to obtain information from sequences and remove errors generated from PCR, such as single nucleotide polymorphisms (SNPs) and chimeric sequences. While many of these are quality approaches, we were unable to find a simple program or workflow with existing programs that provided easily human readable output that detailed variant lineages with the information we wanted and with sufficient removal of chimeric sequences. Specifically, we wished to include deletion and insertion events as well as SNPs and multiple nucleotide polymorphisms (MNP)s in our analysis and be able to view linked variances as single lineages. We also wished to be able to view downstream amino acid changes and have removal of chimeric sequences generated from PCR.

Here we detail the workflow (Fig1) we used to analyze high throughput sequencing data and the program we developed to provide a human readable, information dense output for viewing lineages.

**Figure 1.**
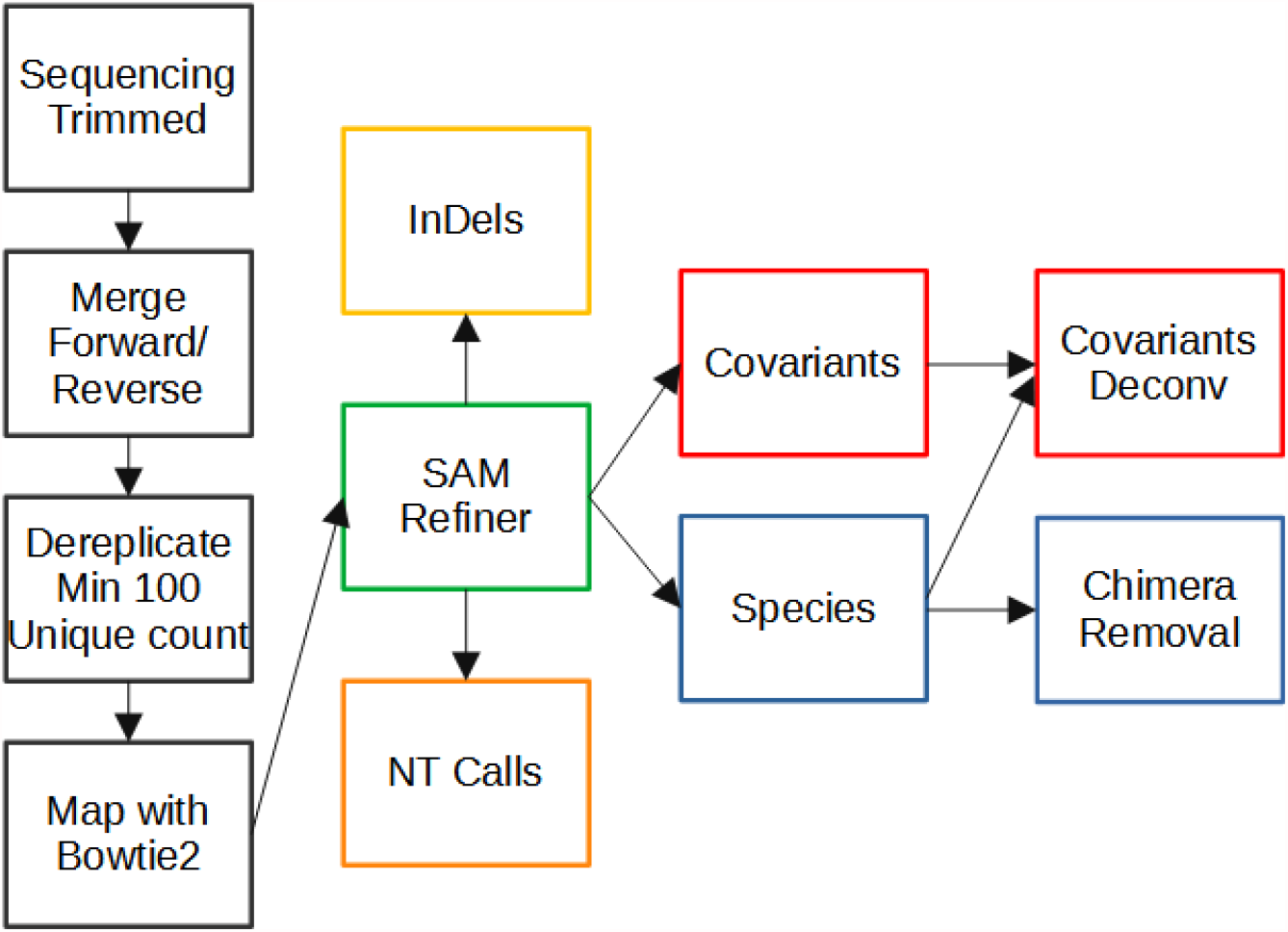
Workflow of Amplicon Sequencing Analysis. Computational processing of sequencing results prior to the use of SAM Refiner is seen in the black boxes. Paired end reads generated from an Illumina MiSeq were trimmed of low quality calls at the end of the reads. Paired end reads were then merged into single contiguous reads. Reads were then dereplicated to unique sequences with at least 100 counts while preserving the count information in the sequence IDs. Dereplicated sequences were then mapped to the sequence of the SARS-CoV-2 Spike ORF using Bowtie2. SAM Refiner was then used to process the mapped reads to obtain information about the variant lineages observed, initially outputting 4 TSV files to report unique sequences, nt calls, indels and covariants. The unique sequences and covariants were further processed to remove chimeric PCR artifacts to produce covariant deconvolution and chimera removed outputs.

## 2. Materials and Methods

### 2.1. Wastewater Collection

Twenty-four hour composite samples were collected at wastewater treatment facilities (WWTF) and were maintained at 4°C until they were delivered to the analysis lab, generally within 24 hours of collection. Samples reported in this study were collected at the NPSD Interim Saline Creek Regional WWTF in Fenton, MO.

### 2.2. RNA Extraction

Wastewater samples were centrifuged at 3,000xg for 10 minutes and then filtered through a 0.22 µM polyethersolfone membrane (Millipore). Approximately 37.5 mL of wastewater was mixed with 12.5 mL solution containing 50% (w/vol) polyethylene glycol 8,000 and 1.2 M NaCl, mixed, and incubated at 4°C for at least 1 hr. Samples were then centrifuged at 12,000xg for 2h at 4°C. Supernatant was decanted and RNA was extracted from the remaining pellet (usually not visible) with the QIAamp Viral RNA Mini Kit (Qiagen) using the manufacturer’s instructions. RNA was extracted in a final volume of 60 µL.

### 2.3. Sequencing

The primary RT-PCR (25 µl) was performed with 5 microliters of RNA extracted from wastewater samples with loci specific primers (0.5 µM each) shown in Table 1 using the Superscript IV One-Step RT-PCR System (Thermo Fisher). Primary RT-PCR amplification was performed as follows: 25°C(2:00) + 50°C(20:00) + 95°C(2:00) + [95°C(0:15) + 55°C(0:30) + 72°C(1:00)] x 25 cycles. Secondary PCR (25 µl) was performed using 5 ul of the primary PCR as template with gene specific primers containing 5’ adapter sequences (0.5 µM each), dNTPs (100 µM each) and Q5 DNA polymerase (NEB). Secondary PCR amplification was performed as follows: 95°C(2:00) + [95°C(0:15) + 55°C(0:30) + 72°C(1:00)] x 20 cycles. A tertiary PCR (50 µl) was performed to add adapter sequences required for Illumina cluster generation with forward and reverse primers (0.2 µM each), dNTPs (200 µM each), and Phusion High-Fidelity DNA Polymerase (1U). PCR amplification was performed as follows: 98°C(3:00) + [98°C(0:15) + 50°C(0:30) + 72°C(0:30)] x 7 cycles +72°C(7:00). Amplified product (10 µl) from each PCR reaction is combined and thoroughly mixed to make a single pool. Pooled amplicons were purified by addition of Axygen AxyPrep MagPCR Clean-up beads in a 1.0 ratio to purify final amplicons. The final amplicon library pool was evaluated using the Agilent Fragment Analyzer automated electrophoresis system, quantified using the Qubit HS dsDNA assay (Invitrogen), and diluted according to Illumina’s standard protocol. The Illumina MiSeq instrument was used to generate paired-end 300 base pair length reads. Adapter sequences were trimmed from output sequences using cutadapt [15]. The raw and trimmed reads for the samples used in this report are available at https://github.com/degregory/SR_manuscript/tree/master/Fenton_Data.

**Table 1.**
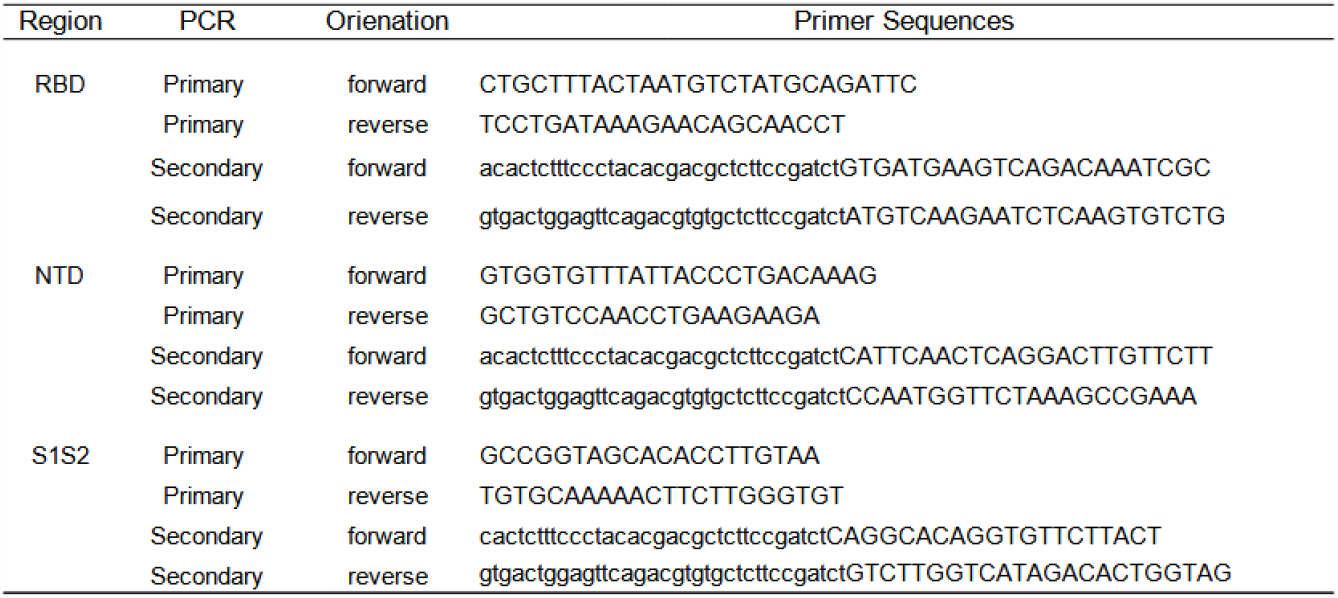
PCR primers used to amplify Spike regions for MiSeq sequencing. Upper case indicate SARS-CoV-2 sequence. Lower case indicates adapter sequence.

## 3. Results

### 3.1. Computational Pre-processing

Figure 1 illustrates the steps of our workflow. The two steps of our process after read trimming used the VSEARCH tool [16]. First, the trimmed paired reads were merged using vsearch – fastq_merge with default parameters. Then merged reads were dereplicated using vsearch -- derep_fulllength with the arguments --minsize 100 and --sizeout. These arguments limit the output to unique sequences that occur at least 100 times and appends the sequence IDs with ‘size=#’, where # is the number of times that sequence occurred in the reads. The cutoff of 100 counts removes late stage PCR errors, leaving only sequences representing the original templates or errors that occurred in early cycles of the PCR. This removal makes further analysis simpler and faster. However, very low frequency original template sequences will also be removed by this cut off, so this step could be skipped to preserve such rare sequences. The resulting unique sequences were mapped to the sequence of SARS-CoV-2 (NCBI Reference Sequence: NC_045512.2, https://www.ncbi.nlm.nih.gov/nuccore/NC_045512) spike ORF using Bowtie2 [17] with default parameters to generate standard SAM formatted files. Having SAM formatted files allows the use of the program we developed for amplicon sequencing results. All files associated with these steps for our analysis of the Fenton, MO sewershed in this manuscript can be accessed at https://github.com/degregory/SR_manuscript/tree/master/Fenton_Data.

### 3.2. SAM Refiner: SAM Processing

Our program, SAM Refiner, is currently a command line based python script and is available at https://github.com/degregory/SAM_Refiner along with updated documentation. In order to run SAM Refiner, a python compiler or interpreter is needed (https://docs.python.org/3/tutorial/interpreter.html). Though only tested in a Linux environment, it should function with other common OSes. Figure 2 shows the command line usage for SAM Refiner. Standard SAM formatted files are the starting point for our program. These files are generated by many mapping programs, such as Bowtie2 [17] or BWA [18]. The default functions of SAM Refiner follow. Files with the extension .sam (case insensitive) in the working directory will be identified and processed. To process SAM files, SAM Refiner must be provided a FASTA formatted file for a reference sequence using the command line argument ‘–r reference.fasta’, where the FASTA file contains the same sequence ID and sequence used to map the sequencing reads in the SAM formatted file. If the IDs of the given reference and the reference of mapped sequences in the SAM file do not match, those sequences will be ignored. If the SAM formatted files were generated from dereplicated or collapsed sequences that still contain the unique read count, SAM Refiner can process the counts from certain formats. SAM Refiner will recognize the counts in sequence ids where the count is at the end of the id and denoted with a ‘=‘ or ‘-’, i.e. ‘Seq1:1;counts=20’ will be recognized as a sequence with 20 occurrences.

**Figure 2.**
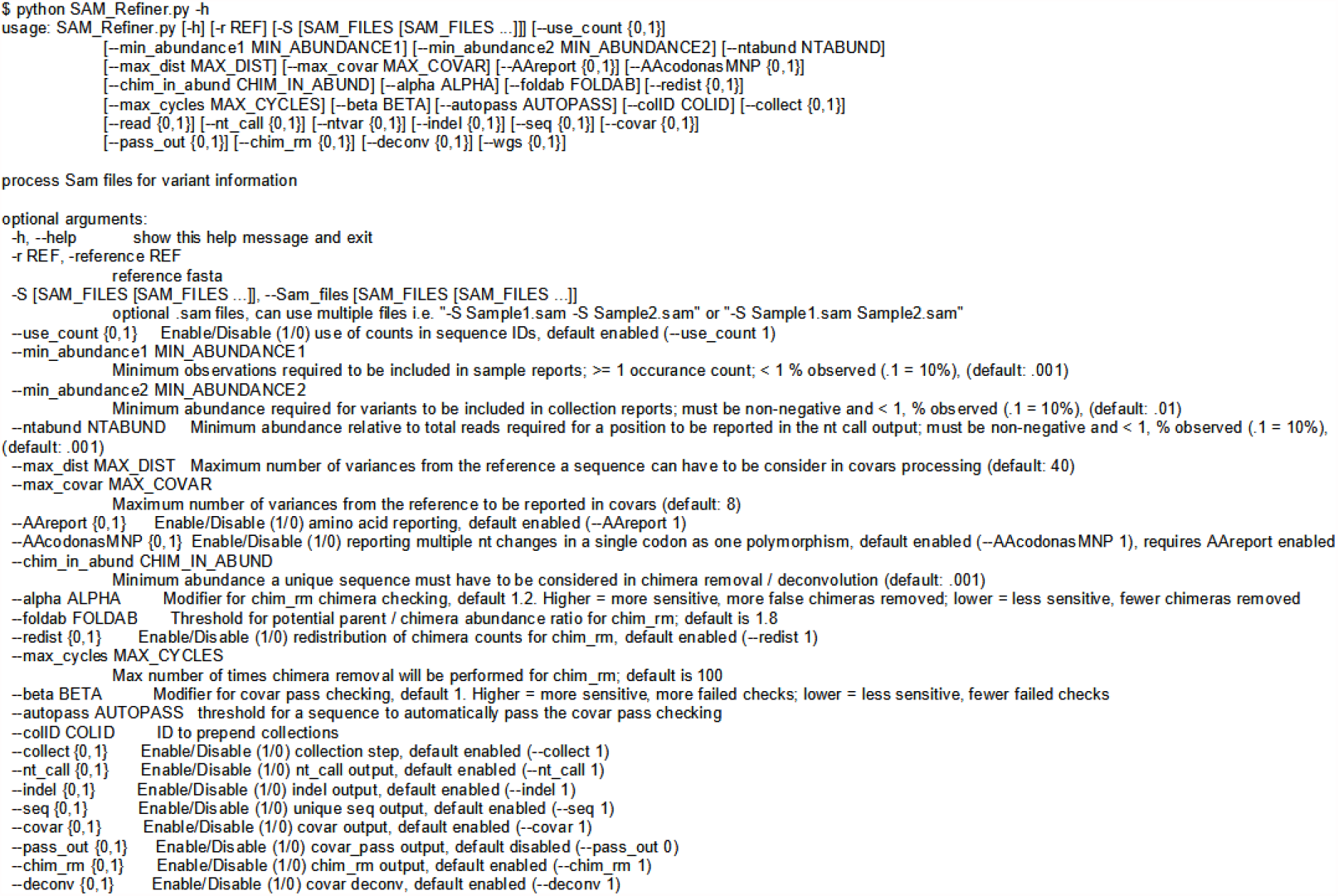
Command Line Usage of SAM Refiner. The standard help output from SAM Refiner is shown. Syntax for the command line usage is seen, followed by details about potential arguments to modify program parameters.

For each SAM file, SAM Refiner initially outputs 4 tab separated values (TSV) files that can be read by any standard spreadsheet software. For a SAM file with the name Sample.sam, the outputs are named Sample_unique_seqs.tsv, Sample_nt_calls.tsv, Sample_indels.tsv and Sample_covars.tsv. Example outputs of each are provided in Supplemental Files 1, 2, 3, and 4, respectively (https://github.com/degregory/SR_manuscript/tree/master/Supplementals). All reports are based on the FASTA reference relative to the SAM formatted file, so any errors made by the mapping or incongruence between the FASTA reference and the mapping reference will result in propagated errors. The reports also include the coded amino acids and their position in the coded peptide as if the reference is an in-frame coding sequence. If multiple nucleotides in a single codon differ from the reference, they will be reported together as a MNP with the associated amino acid change. Within the files, all of the sample specific outputs start with the name of the sample taken from the SAM file name followed in parenthesis by the count of reads mapped.

The Sample_unique_seqs.tsv file (Sup. 1) lists the unique sequence reads mapped in the SAM file using a variance notation to list the variations from the reference along with occurrence count and abundance. For example, using the previously mentioned SARS-CoV-2 spike ORF as the reference sequence, a sequence read that matches the reference except for having a T at position 1501 instead of the reference A would be reported simply as ‘1501A(N501Y)’. The abundance reported uses a decimal notation, so 0.2 represents 20%. Unique sequences that have an abundance below 0.001 are not reported.

The Sample_nt_calls.tsv file (Sup. 2) has a line for each nt position covered in at least 0.1% of the reads. Based on the reference sequence, each line first reports the nt position, the reference nt, the amino acid position, and the reference amino acid residue. The line then reports the number of calls for each base and for deletions at that position, followed by the most abundant (primary) call and its counts and abundance. If that primary nt is different from the reference, the amino acids encoded by the primary nt sequence and by the reference sequence with only that nt changed are reported. Further, if the second (secondary) and third (tertiary) most abundance nts are above .1% of the total read counts, those nts, their counts and abundances, and their associated amino acid changes are also reported.

The Sample_indels.tsv (Sup. 3) file lists each insertion or deletion found in the mapping along with its occurrence count and abundance. Reported insertions have the format of ‘position-insertNT(s)’, so an insertion between nt positions 54 and 55 of the sequence ‘GCA’ will be reported as ‘55-insertGCA’. Reported deletions have the format ‘start Position-end positionDel’, so a deletion of the nts at positions 61 through 64 would be reported as ‘61-64Del’. Amino acid changes are reported if the indel maintains the reading frame. If there are no indels in the reads, no indel report will be generated.

Finally, the Sample_covars.tsv (Sup. 4) file lists all observed single variances and variance combinations relative to the reference sequence. The number and abundance of sequence reads containing each covariant (covar) are reported regardless of whether any of those reads have other variations or not. As an example of this processing, the sequence ‘1212G(G404G) 1501T(N501Y) 1709A(A570D)’ with 100 counts would have the covariants of ‘1212G(G404G)’, ‘1501T(N501Y)’, ‘1709A(A570D)’, ‘1212G(G404G) 1501T(N501Y)’, ‘1212G(G404G) 1709A(A570D)’, ‘1501T(N501Y) 1709A(A570D)’ and ‘1212G(G404G) 1501T(N501Y) 1709A(A570D)’, and contribute 100 counts to each. Because unique sequences that fall below the .1% reporting cutoff can still contribute to covariants, there may be variances in the reported covariants that aren’t seen in the unique sequence output. Any sequence with more than 40 variances from the reference are ignored. While all sequences with 40 or fewer variances are analyzed, only combinations of 8 or less variances are reported.

Once the above outputs are generated from each SAM file found, SAM Refiner will collect information from each sample and report them in a single file for the covars and unique_seqs reports (Collected_Covariances.tsv and Collected_Unique_Seqs.tsv). These collections have a threshold of 1% occurrence for reporting.

Many options are available as command line arguments to change parameters of the SAM processing of SAM Refiner (Fig. 2). There are no strictly required command line arguments, though the -r argument is required for the SAM processing. Omitting the reference will cause SAM Refiner to skip the SAM processing and only perform the collections and chimera removal (see below), which require per-exisiting outputs. The other input option is the ‘-S’ argument which provides SAM Refiner with SAM files to process instead of allowing it to search the working directory. The use of dereplicated/collapsed counts in the SAM files can be disabled with ‘--use_counts 0’. There are also options for the outputs. All outputs can be separately suppressed with the arguments ‘--seq 0’, ‘--nt_call 0’, ‘--indel 0’, ‘--covar 0’ and ‘--collect 0’. The collections file names can be prepended with a string specified by the argument ‘--colID’. To change the reporting threshold for the sample and collected outputs, arguments ‘--min_abundance1’ and ‘--min_abundance2’ are used respectively. For ‘--min_abundance1’, despite its name, the value can be used to either set a minimal abundance threshold or a minimal count threshold. Values of 1 or greater will set a count threshold, while those less than 1 will set an abundance threshold. Only an abundance threshold is available for ‘--min_abundance2’. All amino acid information in the reports can be suppressed with the argument ‘--AAreport 0’. This disabling is recommended if the reference doesn’t primarily provide an in frame coding sequence. Users can also have all nt changes processed independently, even if they are in the same codon with --AAcodonasMNP 0’.

Using ‘--ntabund’ will change the required mapped coverage threshold for reporting a position in the nt_calls output. Finally, ‘--max_dist’ and ‘--max_covar’ allow changes to the covar processing and reporting. Sequences with more variations than the amount specified by ‘--max_dist’ are not included in the covar analysis. The maximum number of variances reported in a combination can be set with ‘--max_covar’. As an example, if ‘--max_covar 2’ were used for Sup. 4, then ‘1216-1216Del 1501T(N501Y) 1709A(A570D)’, ‘1212G(G404G) 1501T(N501Y) 1709A(A570D)’ and ‘1217-1217Del 1501T(N501Y) 1709A(A570D)’ would not be reported.

Using the SAM files generated from the sequencing data of the Fenton sewershed, we ran SAM Refiner with the SARS-CoV-2 (NCBI Reference Sequence: NC_045512.2) spike ORF sequence as a reference, the same as was used with the Bowtie2 mapping. The resulting outputs can be accessed at https://github.com/degregory/SR_manuscript/tree/master/Fenton_Data. These outputs allow us to see the variant lineages present at different dates in this sewer shed. However, as can be seen in Sup. 1, many of the sequences reported appear to be chimeric sequences arising from template jumping. While these outputs can still be used for further analysis, removing chimeric sequences makes such analysis easier. SAM Refiner also has methods to remove such chimeric sequences.

### 3.3. SAM Refiner: Chimera Removal

PCR amplification can introduce sequence errors that obscure the original template sequences. Of most concern are the introduction of false SNPs and chimeric reads. Most PCR introduced SNPs can be removed from analysis by the use of an abundance threshold such as is the default for SAM Refiner, or as was used in our pre-processing dereplication step. There are also numerous programs that can be used to attempt to remove such errors. Chimeric sequences are generally more difficult to remove. Many programs exist for this task; however, we were unable to find any that provided satisfying results for our amplicon sequencing. We developed two algorithms for SAM Refiner in order to remove chimeric errors arising from PCR template jumping from the SAM processing outputs. They are redundant in their function but use different methods, allowing for increased confidence in results by crosschecking between the two methods.

The algorithms to remove chimeric sequences rely on the unique sequence and covariant files generated by the SAM processing. The first algorithm, chimera removed (chim rm), goes through the individual unique sequences, starting with the lowest abundance, and determines if the sequences are chimeric. Figure 3 shows a simplified example of how the determination is made on the lowest abundant sequence of an example unique sequence output (Sup. 5). The more detailed and exact method is as follows. The sequence being considered as a potential chimera is broken up into all possible dimeric halves. Each pair is then compared to all the other sequences to detect potential parents. A sequence is flagged as a potential parent if its abundance is greater than or equal to the abundance of the potential chimera multiplied by 1.8 (foldab) and there is at least one other sequence that would be a matched parent to the complimentary dimeric half. When a pair of dimeric halves have potential parents, the abundances of parent pairs are multiplied. The products from each potential parent pairings are summed as an expected abundance value and compared to the observed abundance of the potential chimera. If the potential chimeras abundance is less than that of the expected value multiplied by 1.2 (alpha), that sequence is flagged as a chimera and removed. The counts attributed to that flagged chimeric sequence are then redistributed to the parents sequences based on the relative expected contribution to recombination. Once this process has been done for all the sequences, it is repeated until no more sequences are flagged as chimeric or 100 chimera removal cycles have completed. The results of this algorithm that have a recalculated abundance of 0.001 or greater are output in a new file (Sup. 6 Example_a1.2f1.8rd1_chim_rm.tsv). The added string represent values of the parameters used for the processing (alpha, foldab and redist, see below for more information on the parameters).

**Figure 3.**
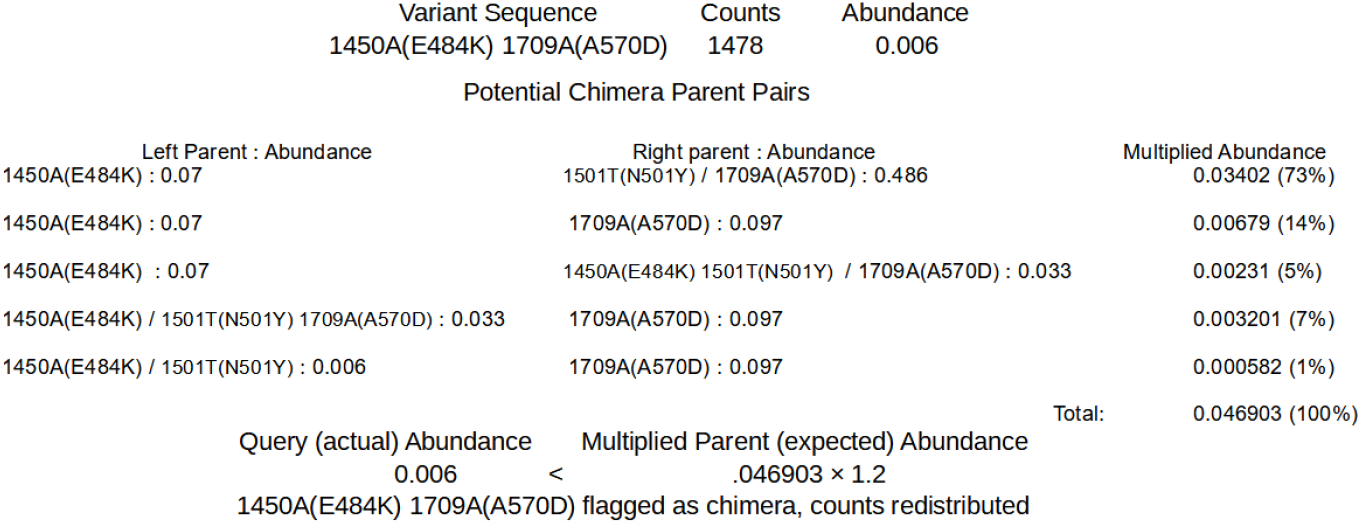
First method of detection and removal of chimeras, Chimera Removed. Using the sequences shown in Sup. 5, the query of the least abundant sequence is shown. Potential parents whose recombination could result in the query sequence are found. The abundances of each potential pair are multiplied. The sum the multiplied abundances of the pairs (expected) is then compared to the abundance of the query sequence (actual) to determine if the query sequence is a chimera. If the actual abundance is greater or equal to 1.2 times the expected abundance, the sequence is considered non-chimeric.

The second algorithm, covariant deconvolution (covar deconv), is a two-step process. Figure 4 shows these processes using example outputs in Sup. 5 and 7. The first step determines if a sequence is likely to be a true or chimeric sequence by obtaining the ratio of the frequency of a given covariant sequence relative to an expected abundance of that covariant sequence assuming random recombination of its individual polymorphisms. The expected abundance is obtained by multiplying the abundances of each individual variance that is present in that covariant sequence. For instance, in a sample where ‘1501T(N501Y)’ has an abundance of 0.32 and ‘1709A(A570D)’ has an abundance of 0.35, the expected abundance of the covariant ‘1501T(N501Y) 1709A(A570D)’ would be 0.112 [0.32 × 0.35]. If the ratio of the observed abundance to the expected abundance is equal to or greater than 1 (beta), that covariant passes the check and is sent to the second step. Any sequence that has an abundance of 0.3 or greater is automatically passed. If such a sequence has an observed/expected ratio less than 1, it will be assigned a ratio of 1. The second step processes the passed sequences in order of greatest observed/expected ratio to least. If multiple sequences have the same ratio, they are processed in order of greatest to least distance from the reference. Sequences that automatically pass the first step are processed after the other sequences and in order of least abundant to greatest. Sequences are assigned a new occurrence count based on their constituent individual variances. For the sequence being processed, the count for the least abundant individual variance is assigned to the sequence and constituent variances making up the sequence have their count reduced by the amount of the least variance. This reduction means the individual variance that had the least counts is assigned 0 counts, so any sequence not yet processed in which that variance is present is functionally removed. This process is repeated until all sequences have been reassessed or removed. The final results with an abundance of 0.001 or greater are reported in a new file (Sup. 8 Example_covar_deconv.tsv).

**Figure 4.**
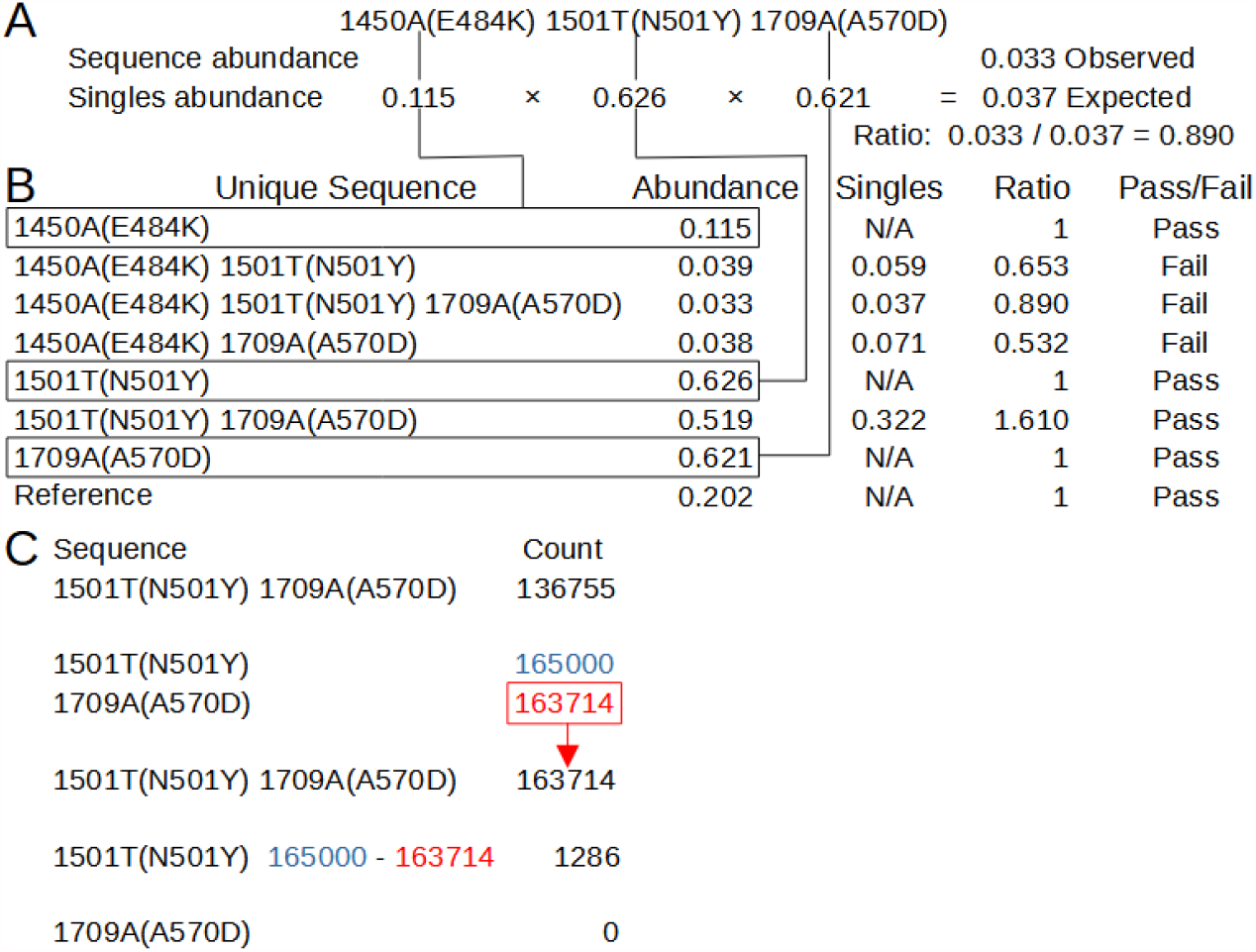
Second chimera removal method in SAM Refiner, Covariant Deconvolution. A. Calculations of the singles / expected abundance and abundance ratio for one of the unique sequences from Sup 5 and the abundances from Sup 7. Lines connect the singles and their abundance to the same in B. B. Calculations for determining if a unique sequence passes the initial check. Sequences pass when they have an Abundance/Singles ratio of 1 or greater. C. Passed sequences are processed in order of greatest ratio to least. Counts of the sequence are set to the counts of the least abundant single variant, and that count is then removed from all single variants in that sequence.

As before, the results from individual samples are collected and reported for entries above 1% occurrence. A number of command line arguments will also influence the chimera removal algorithms. Both chimera removal algorithms run by default, but either or both can be disabled (--chim_rm 0 and –covar_deconv 0). The collections are again disabled with ‘--collect 0’. An additional output of the covariants that passed the first step of the second algorithm can be generated with ‘--pass_out 1’(Sup. 9). The outputs are constrained as before by a minimum abundance with command line arguments ‘--min_abundance1’ and ‘--min_abundance2’. Collection file names are also prepended with ‘--colID’. The only input parameter that can be changed by command line argument is the abundance of sequences or covariants that will be considered in the algorithms. By default, only entries from the inputs that have a 0.001 abundance or greater are processed. This threshold can be changed with ‘--chim_in_abund’.

Four parameters can be altered for the first algorithm. The abundance ratio that is used as a threshold for selecting potential parents of potential chimera can be set with ‘--foldab’. Larger values will generally reduce the pool of sequences that will be considered as potential parents, thus potentially reducing the total expected abundance obtained from parent pairs and number of sequences flagged as chimeric. In the most simple theoretical model of PCR chimera generation, two parents generate one chimera. The parents have at least twice the abundance of the chimera as they would exist and have been amplified prior to the chimera. The reality of chimera generation can be much more complex, as many sequences may generate identical chimeras multiple times. If a sample has little chimera generation, a --foldab value close to 2, such as the default of 1.8, should be sufficient to remove chimeras without also removing non-chimeric sequences in error. However, the more chimera generation observed, the more the --foldab value needs to be reduced to accurately remove all chimeric sequences, even to 0 to not exclude any sequence from being considered a potential parent (though it will likely be vary rare for such a value to be necessary). Lower values, however, will also increase the likelyhood of a sequence being flagged as a chimera in error. Users may need to empirically determine the best value for their samples.

The multiplier for the parental summed abundance for determining if a sequence is a chimera can be set with ‘--alpha’. Larger values will generally result in a greater number of sequences flagged as chimeric. As with --foldab, the optimal value for --alpha will depend on the extent of chimera generation in the samples being processed, with a value near 1 for minimal chimera generation (such as the default 1.2) and 2 or even higher for rampant chimera generation. Once again, the later would also increase the likelihood of sequences being flagged as chimeric in error.

Redistribution of the counts from the chimera to the parent sequences can be disabled with ‘--redist 0’. Redistribution is meant to give an estimate of the counts and abundances that would have been observed without chimera generation, which users may wish to forgo. The maximum number of chimera removal cycles can be change by ‘--max_cycles’, ei ‘--max_cycles 2’ will only allow two iterations of the chimera removal. Multiple removal cycles allows chimeras to be found based on new counts and abundances resulting from previous cycles, increasing the likelihood chimeras are removed from a sample.

The second algorithm has two parameter that can be changed. The ratio threshold at which a covariant will be passed to the second step can be altered with ‘--beta’. The abundance at which a covariant will automatically be passed can be changed with ‘--autopass’.

The chimera removal methods of SAM Refiner were also used on the Fenton sewershed sequencing data. Due to the relatively high amount of chimeric sequences in our samples, we used the command line arguments ‘--foldab=0.6 –alpha=2.2’. The outputs generated for the Fenton sewershed from 2-2-21 to 4-13-21 can be accessed at https://github.com/degregory/SR_manuscript/tree/master/Fenton_Data. The two different chimera removal methods showed good concordance, validating each as being a viable method. Duplicate RT-PCR preparation and sequencing of the same wastewater sample also generally provided similar results, though less consistently (Fig. 5. Compare A and B RBD amplicon preparations). These differences were more pronounced with covariants with relatively low abundance, such as is seen with 3-30 RBD samples, where one detects T478K and the other does not (Fig. 5). These differences illustrate the stochastic nature of RT-PCR amplification.

**Figure 5.**
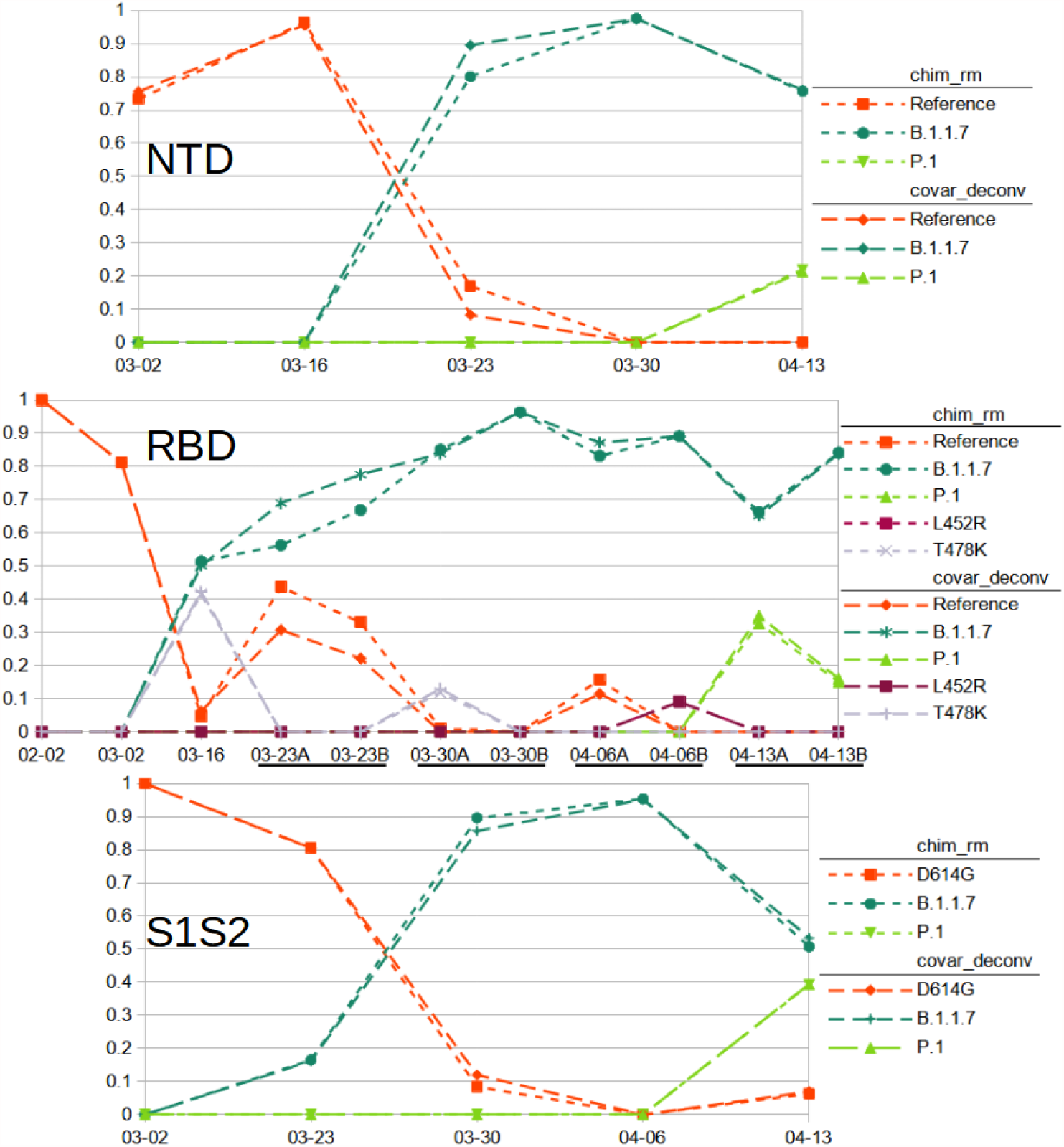
Relative Abundance of Reference and Variant SARS-CoV-2 Sequences Observed in Fenton, MO sewershed from February to March. Results from sequencing of Spike amplicons of the NTD, RBD and S1S2 junction regions are shown. Lines of short dashes connect values obtained by the chimera removed method, lines of long dashes connect values obtained by the covariant deconvolution method. All amplicons show a population shift from the reference with D614G to B.1.1.7 sequences with the appearance of P.1 sequences at the last time point. Additionally, known common polymorphisms T478K and L452R were observed from the RBD amplicons. RT-PCR for the RBD amplicon was performed in duplicate for some samples.

We used the chimera removed and covariant deconvolution outputs to assign sequences to known variant lineages or the reference (Sup. 10, 11 & 12) based on variances present. Variances that only appeared in one sequencing run and did not appear frequently in GSIAD (https://www.gisaid.org/) were considered likely PCR error and not taken into account for sequence assignment. Based on the assignments, we were able to observe the changes to virus populations in the sewershed over time (Fig. 5). We classified the sequences found from the NTD amplicon as matching reference sequence, lineage B.1.1.7 with ‘203-208Del 429-431Del’ or lineage P.1 with ‘412T(D138Y) 570T(R190S)’ (Sup 10). Sequences from the RBD amplicon matched reference sequence, lineages B.1.1.7 with ‘1501T(N501Y) 1709A(A570D)’ or P.1 with ‘1250C(K417T) 1450A(E484K) 1501T(N501Y)’, or had the single variations of T478K or L452R (Sup 11). T478K and L452R each have lineage associations. However, no other variances are associated with these in the RBD amplicons, nor were any variances present in the other amplicons that would indicate the presence of any associated lineages. While these SNPs could be the result of PCR error, it is more likely the associated lineages exist in the sewershed, but due to stochastic effects the other associated variances in the other amplicons were not detected. They could have also arisen in the reference background. As we can not assign them to known variant lineages with any certainty, we assigned them to their own category. Sequences from the S1S2 amplicon matched lineage B.1.1.7 with ‘1841G(D614G) 2042A(P681H) 2147T(T716I)’, lineage P.1 with ‘1841G(D614G) 1963T(H655Y) 2063T(A688V)’ or only had the now ubiquitous D614G variation (Sup 12). The 03-23 S1S2 sample had a sequence ‘1841G(D614G) 2037G(N679K) 2063T(A688V)’. While A688V is associated with P.1, it does not appear in that context here. As that is the only sample where those covariant sequences were observed and the variances are not frequently reported in GISAID (outside of P.1 for A688V), we assigned it to the reference category. Looking at all samples over time and the three amplicon regions in concert, we can conclude that the SARS-CoV-2 population of this sewershed changed from almost exclusively having only the D614G variation to mainly the B.1.1.7 lineage, with the introduction of P.1 early in April. This general method is now being used to track SARS-CoV-2 variants in many Missouri sewersheds (https://storymaps.arcgis.com/stories/f7f5492486114da6b5d6fdc07f81aacf).

### 3.4. SAM Refiner: Limitations and Future Development

While the outputs of SAM Refiner can be very informative, the program has some limitations, some of which may be overcome in future development. Currently the greatest limitation is the need for users to be familiar with command line usage. We hope to develop a graphical user interface version of these programs to overcome this user hurdle in the future. We also intend to develop SAM Refiner to be available from widely used functional collections, such as BioConda (https://bioconda.github.io/) and Galaxy (https://usegalaxy.org/).

Though SAM Refiner can be used on sequencing not based on amplicons, its usefulness will be more limited, as the relative abundance of sequences and covariants will be calculated based on total reads and not positional coverage. Future development may include modes for whole genome sequencing or multiple amplicons, even the ability to use multiple sequences for a reference.

The accuracies of the chimera removal algorithms will vary greatly depending on the parameters used and the sample they are being run on. Due to the stochastic nature of chimera generation and amplification during PCR and the possible complexity of the original template sequences, samples will sometimes be refractory to chimera removal algorithms. This problem is faced by all programs designed for this purpose. The ability to modify parameters in the algorithms and having two algorithms with different approaches to the chimera removal improves the accuracy the user can achieve with this software. Some samples will, however, always fail to be processed accurately by one or both methods.

## Data Availability

All data is available at https://github.com/degregory/SR_manuscript

## Supplementary Materials

The following are available online at https://github.com/degregory/SR_manuscript/tree/master/Supplementals, Sup. 1 Example of SAM Refiner’s Output for Reporting Unique Sequences, Sup. 2 Example of SAM Refiner’s Output for Reporting Positional NT Calls, Sup. 3 Example of SAM Refiner’s Output for Reporting Insertions and Deletions, Sup. 4 Example of SAM Refiner’s Output for Reporting Covariance, Sup. 5 Sample Unique Sequences Output With Chimeric Sequences, Sup. 6 Sample Output of Sequences of SAM Refiner’s Chimeras Removed, Sup. 7 Sample Covariance Output With Chimeric Sequences, Sup. 8 Sample Passed Sequences Output from the First Part of SAM Refiner’s Covariant Deconvolution Method, Sup. 9 Sample Output of Sequences by SAM Refiner’s Covariant Deconvolution Method, Sup. 10 Assignment of NTD Covariant Sequences to Variants and Lineages, Sup. 11 Assignment of RBD Covariant Sequences to Variants and Lineages, Sup. 12 Assignment of S1S2 Covariant Sequences to Variants and Lineages

## Author Contributions

Conceptualization, MJ.; methodology, MJ; software, DG; validation, DG and MJ.; formal analysis, DG and MJ; investigation, DG and MJ; resources, MJ, JW; data curation, DG, CW, CL and MJ; writing—original draft preparation, DG; writing—review and editing, DG, JW, CW and MJ; visualization, DG; supervision, MJ; project administration, CW, JW, and MJ; funding acquisition, JW and MJ. All authors have read and agreed to the published version of the manuscript

## Data Availability Statement

Raw and processed data can be accessed at https://github.com/degregory/SR_manuscript

## Acknowledgments

We would like to acknowledge Christopher Bottoms for assistance in software development, and the University of Missouri DNA Core for assistance in developing deep sequencing protocols.

